# Retrospective Characterization and Review of 466 New Daily Persistent Headache Cases Within a Quaternary Healthcare System

**DOI:** 10.1101/2025.11.26.25341079

**Authors:** Zeshan Tariq, John Majewski, Bridget Lloyd, Nicky Don, Viraj Govani, Alfred Vinnett, Olivia Babich, Selina J. Chang, Alison Choi, Faith Kehinde, Arjun Mittal, Robert Kaniecki

**Affiliations:** Department of Neurology, University of Pittsburgh School of Medicine, UPMC Headache Center, 120 Lytton Ave, Suite 300, Pittsburgh, PA, 15213, USA; University of Maryland School of Medicine, 655 West Baltimore Street, Baltimore, MD, 21201, USA

**Keywords:** New Daily Persistent Headache, NDPH, Chronic Daily Headache, Primary headache disorders

## Abstract

**Background:** New Daily Persistent Headache (NDPH) is an enigmatic syndrome characterized by a distinct onset headache that becomes continuous and unremitting within 24 hours and persists for at least 3 months without underlying structural cause. Patients may experience years or decades of pain with negative impacts on functionality and quality of life. Although characteristically treatment resistant, case reports of response to pharmacologic therapies exist, though with great heterogeneity and without generalizability. Given NDPH is incompletely characterized and displays phenotypic variability, it is possible that better sub-population identification might yield more targeted therapeutic directions.

**Methods:** We conducted a retrospective chart review of adult patients seen within the University of Pittsburgh Medical Center System who received an NDPH International Classification of Diseases 9 or 10 code between 2011-2019. 5,926 patient charts were reviewed and only those who satisfied International Classification of Headache Disorders, 3^rd^ edition criterion were further analyzed. Two independent data collectors reviewed each chart.

**Results:** 466 individuals satisfied NDPH diagnostic criteria. Median age at first consultation was 37.1 years, with 66.7% of the population identifying as female. Length of NDPH duration at the last consultation was 1.75 years. Headache onset exhibited no obvious seasonality. Headaches were unilateral in 12.5% of cases and severe pain was reported in 79.1% of cases with documented pain severity. Thunderclap onset was observed in 5.8% of cases. Further categorization into migraine (MP) and tension (TP) phenotypes found approximately even distribution, 207 and 214 respectively. Both MP and TP exhibited demographics, precipitating factors, and prognostic types reflective of the general population. In comparison to TP, MP patients described increased prevalence of severe pain (48.3% vs 14.0%), nausea (68.6% vs 33.6%), vomiting (25.1% vs 7.0%), light-sensitivity (85% vs 27.1%), noise-sensitivity (80.7% vs 24.8%), and osmophobia (19.3% vs 3.7%). Comparison of MP and TP exacerbating factors found increased reporting of physical activity (23.7% vs 10.3%). Chronicity of MP and TP found similar breakdowns with 26.1% vs 23.4% reporting 5+ years of headache.

**Conclusions:** NDPH is a refractory, severe pain headache disorder that exhibits no seasonality for onset, with equal breakdown of MP and TP subtypes.

## Background

New Daily Persistent Headache (NDPH) is a physiologically enigmatic syndrome resulting in a classically treatment-resistant, unremitting headache without an underlying structural cause.^1^ ICHD-3 criteria describe NDPH as a primary headache disorder of distinct and clearly remembered onset, with pain becoming continuous and unremitting within 24 hours and persisting for >3 months.^2^ Patients may experience years or decades of pain with negative impacts on functionality and quality of life.^1^ Although most patients are unresponsive to therapeutic interventions, case reports have demonstrated therapeutic benefit with venlafaxine,^1,3^ nimodipine^4^, intravenous methylprednisone,^5^ onabotulinum toxin type A,^6^ and osteopathic manipulation,^7^ which suggest certain patients may respond to treatment. Yet the wide heterogeneity in responsiveness to these treatments suggests a potential application for NDPH subtyping. One option is to subtype subjects based on the nature of onset. For example, several NDPH patients who specifically endorsed a thunderclap onset exhibited responsiveness to nimodipine.^4^ Another possible feature for subtyping that remains less explored is reported potential triggering events, such as a stressful life event, viral infection,^8^ or surgical procedure.^9^

Given the potential benefit in subtyping, there is an obvious impetus to characterize and expand our understanding of the NDPH disease landscape. To date, there are relatively few such studies, with most having limited case sample sizes due, in part, to the low overall prevalence rate of NDPH.^10^ Published data typically includes dozens or several hundred cases: 56 USA;^1^ 30 Japan;^11^ 18 Italy;^12^ 71 USA;^13^ 92 Taiwan^14^; 63 India;^15^ 38 China;^16^ 55 India;^17^ 162 UK;^18^ 97 USA;^19^ 76 UK;^20^ 22 USA;^20^ 328 USA.^21^ Furthermore, the heterogeneous methodology of these studies makes it difficult to conduct cross comparisons limiting signal to noise differentiation.

To address these limitations in the existing literature, we conducted a large-scale retrospective chart review of all patients who received an International Classification of Diseases (ICD) 10 (’G44.52’) or ICD 9 (’339.42’) diagnostic code within the University of Pittsburgh, Medical Center System (UPMC) from 2011 to 2019, ultimately collecting information on 466 NDPH patients while mimicking study design from one of the prior largest NDPH chart review studies for comparison purposes.^21^

## Methods

### Methodology of Chart Review

All adult patients seen at UPMC between 2011 and 2019 who received a diagnostic code of either ICD 10 (’G44.52’) or ICD 9 (’339.42’) were collated into a database for manual screening. In total, 5,926 patient charts were reviewed, and only subjects who satisfied NDPH criterion as defined by International Classification of Headache Disorders, 3rd edition (ICHD-3) were further analyzed. Two independent data collectors reviewed, each chart with any disagreements resolved by a third reviewer with guidance being provided by senior author R.K., a neurologist with subspeciality certification in Headache Medicine.

### Statistical analysis

Descriptive statistics included frequencies and percentages. Means and standard deviations (SD) were computed for normally distributed data, and medians [25th, 75th] were computed for non-normally distributed data. Standardized mean differences (SMD), the between-group differences divided by in-group variability (also known as Cohen’s D), were computed for migraine and tension phenotypes based on demographics, onset month, precipitating factors, clinical features, exacerbating factors, and prognostic types. If a variable was a “sub-level” of a larger variable, that is, a patient could only exclusively select one of the possible options (e.g. “mild,” “moderate,” or “severe” for quality of initial pain), then only one standardized mean difference was calculated. Missing values were included in the calculation of the SMD and overall percentages, but not depicted as separate columns in tables, except for the “unknown” category for “Month of Onset,” and certain demographic variables as noted.

All analyses were conducted in R 4.4.1 (2024-06-14), R 4.3.1 (2023-06-16), R 4.3.0 (2023-03), R Studio 2023.06.0+421, and Posit Cloud (an online R platform), and relied on CreateTableOne SMD calculations. The sample size was based on the available data. No statistical power calculation was conducted.

### Ethical Approval

The University of Pittsburgh Institutional Review Board provided ethical approval and determined that the study was exempt and granted a waiver of HIPAA authorization.

## Results

A total of 5296 patient charts were reviewed for potential inclusion in the study. Of these 466 individuals satisfied ICHD-3 criteria for NDPH. Reasons for exclusion included the presence of structural secondary causes, noncontinuous headache, a diagnosis of medication overuse headache, or the absence of sufficient historical information to confirm the diagnosis of NDPH.

The demographics of all 466 patients are shown in Table 1. Mean age of onset was 37.1 years, with 66.7% of this population identifying as female and 85.8 % identifying as white. Median duration of NDPH at the time of the last physician appointment was 1.75 years. In 12% of cases, the headaches were strictly unilateral. The month of headache onset varied, with January (8.6%), April (7.1%), September (7.3%), and October (7.5%) having the highest percentages of NDPH onset, while May (3.6%), June (3.9%), and August (3.4%), had the lowest percentages. Prior instances of episodic headaches, potentially representing migraine, were reported by 10.3% of patients.

**Table 1.** Demographics of All NDPH Patients.

| Demographic | All Patients<br>(n=466) | Female<br>(n=311) | Male<br>(n=143) |
| --- | --- | --- | --- |
| Age at onset, mean (SD) | 37.1 (18.6) | 34.6 (18.6) | 43.0 (17.4) |
| Onset month, n (%) |  |  |  |
| January | 40 (8.6) | 26 (8.4) | 14 (9.8) |
| February | 24 (5.2) | 17 (5.5) | 7 (4.9) |
| March | 20 (4.3) | 17 (5.5) | 3 (2.1) |
| April | 33 (7.1) | 22 (7.1) | 11 (7.7) |
| May | 17 (3.6) | 14 (4.5) | 3 (2.1) |
| June | 18 (3.9) | 11 (3.5) | 7 (4.9) |
| July | 23 (4.9) | 16 (5.1) | 7 (4.9) |
| August | 16 (3.4) | 11 (3.5) | 5 (3.5) |
| September | 34 (7.3) | 19 (6.1) | 15 (10.5) |
| October | 35 (7.5) | 20 (6.4) | 15 (10.5) |
| November | 24 (5.2) | 17 (5.5) | 7 (4.9) |
| December | 23 (4.9) | 15 (4.8) | 8 (5.6) |
| Unknown | 159 (34.1) | 106 (34.1) | 41 (28.7) |
| Ethnicity, n (%) |  |  |  |
| Asian | 5 (1.1) | 3 (1.0) | 2 (1.4) |
| Black | 35 (7.5) | 28 (9.0) | 7 (4.9) |
| Hispanic | 3 (0.6) | 2 (0.6) | 1 (0.7) |
| White | 400 (85.8) | 274 (88.1) | 126 (88.1) |
| Other | 3 (0.6) | 3 (1.0) | 0 (0.0) |
| Unknown | 20 (4.29) | 1 (0.3) | 7 (4.9) |
| Duration of NDPH at last visit (years), median [25th, 75th] | 1.75 [0.58, 5] | 2.0 [0.58, 5] | 1.3 [0.58, 3.5] |
| History of migraine, n (%) | 48 (10.3) | 37 (11.9) | 11 (7.7) |
| Initial Lateralization of Pain, n (%) |  |  |  |
| Unilateral (*Left or Right) | 56 (12.0) | 37 (11.9) | 19 (13.3) |
| Bilateral | 276 (59.2) | 183 (58.8) | 93 (65.0) |
| Left | 34 (7.3) | 24 (7.7) | 10 (7.0) |
| Right | 22 (4.7) | 13 (4.2) | 9 (6.3) |
| Unknown | 134 (28.8) | 91 (29.3) | 31 (21.7) |
| Final Lateralization of Pain (%) |  |  |  |
| Unilateral (*Left or Right) | 46 (9.9) | 32 (10.3) | 14 (9.8) |
| Bilateral | 318 (68.2) | 222 (71.4) | 96 (67.1) |
| Left | 27 (5.8) | 19 (6.1) | 8 (5.6) |
| Right | 19 (4.1) | 13 (4.2) | 6 (4.2) |
| Unknown | 102 (21.9) | 57 (18.3) | 33 (23.1) |

For further analysis, patients were subtyped into migraine (MP) and tension-type phenotypes (TP), with their demographics being reported in Table 2. 207 MP and 214 TP NDPH patients were identified with mean onset ages of 34.7 and 39.2 years old, respectively. Both phenotypes exhibited equivocal patterns of onset months, with similarities in racial/ethnic makeup, duration of NDPH, and pain lateralization. There was a greater history of comorbid migraine headache within the MP compared to TP (15% and 6.1%, respectively). Precipitating factors for both MP and TP are reported in Table 3 and show similar distributions between cohorts. Overall, infection was the most commonly reported precipitating factor (11.4%).

**Table 2.** Demographics by NDPH Type.

| Demographic | Migraine-Type NDPH<br>Patients Seen (%)<br>n=207 | Tension-Type NDPH<br>Patients Seen (%)<br>n=214 |
| --- | --- | --- |
| Age at onset, mean (SD) | 34.7(17.1) | 39.2 (20.1) |
| Onset month, n (%) |  |  |
| January | 19 (9.2) | 17 (7.9) |
| February | 14 (6.8) | 9 (4.2) |
| March | 12 (5.8) | 7 (3.3) |
| April | 11 (5.3) | 20 (9.3) |
| May | 6 (2.9) | 8 (3.7) |
| June | 11 (5.3) | 6 (2.8) |
| July | 7 (3.4) | 13 (6.1) |
| August | 8 (3.9) | 8 (3.7) |
| September | 20 (9.7) | 14 (6.5) |
| October | 14 (6.8) | 20 (9.3) |
| November | 12 (5.8) | 9 (4.2) |
| December | 9 (4.3) | 9 (4.2) |
| Unknown | 64 (30.9) | 74 (34.6) |
| Race/ethnicity, n (%) |  |  |
| Asian | 2 (1.0) | 3 (1.4) |
| Black | 22 (10.6) | 10 (4.7) |
| Hispanic | 2 ( 1.0) | 1 (0.5) |
| White | 173(83.6) | 186 (86.9) |
| Unknown | 3 (1.4) | 0 (0.0) |
| Duration of NDPH at last visit (years),<br>median [25th, 75th] | 1.63 [0.58, 4.73] | 1.75 [0.58, 5.00] |
| History of migraine, n (%) | 31 (15.0) | 13 ( 6.1) |
| Initial Lateralization of Pain, n (%) |  |  |
| Unilateral (Left or Right) | 30 (14.5) | 22 (10.2) |
| Bilateral | 120 (58.0) | 138 (64.5) |
| Left | 16 (7.7) | 14 (6.5) |
| Right | 14 (6.8) | 8 (3.7) |
| Unknown | 57 (27.5) | 54 (25.2) |

| Final Lateralization of Pain (%) |  |  |
| --- | --- | --- |
| Unilateral (Left or Right) | 28 (13.5) | 15 (7.0) |
| Bilateral | 148 (71.5) | 137 (64.0) |
| Left | 13 (6.3) | 11 (5.1) |
| Right | 15 (7.2) | 4 (1.9) |
| Unknown | 31 (15.0) | 62 (29.0) |

**Table 3.** Precipitating Factors.

| Precipitating Factors | Migraine Phenotype Patients Seen n=207 (%) | Tension Phenotype Patients Seen n=214 (%) | Total Number of Patients Seen n= 466 (%) | Standardized Mean Difference (SMD) |
| --- | --- | --- | --- | --- |
| Stressful Life Event | 8 (3.9) | 11 (5.1) | 19 (4.1) | 0.067 |
| Infection | 26 (12.6) | 22 (10.3) | 53 (11.4) | 0.08 |
| Surgery | 8 (3.9) | 14 (6.5) | 28 (6.0) | 0.123 |

Table 4 details clinical features for MP and TP such as the thunderclap onset, nature of the pain (throbbing, pressure, aching, other), intensity upon initial consult (mild, medium, severe), laterality, and concomitant characteristics (nausea, vertigo, dizziness, vomiting, light sensitivity, noise sensitivity, visual aura, tinnitus onset, neck pain), accompanied by an equivalent summary for the total NDPH patient cohort. Using a visual analog scale of 1 (mild) to 10 (severe), pain was rated as follows: mild, 1-3; moderate, 4-6; and severe, 7-10. MP had greater association with throbbing and increased pain severity compared to TP. Additionally, nausea, dizziness, vomiting, light sensitivity, noise sensitivity, visual aura, and osmophobia were more often present in MP.

**Table 4.** Clinical Features.

| Clinical Features | Migraine Phenotype<br>Patients Seen<br>n=207 (%) | Tension Phenotype<br>Patients Seen<br>n=214 (%) | Total Number of<br>Patients Seen<br>n= 466 (%) | SMD |
| --- | --- | --- | --- | --- |
| Thunderclap Onset | 14 (6.8) | 8 (3.7) | 27 (5.8) | 0.375 |
| Quality of Pain |  |  |  |  |
| Throbbing | 89 (43.0) | 48 (22.4) | 144 (30.9) | 0.465 |
| Pressure | 61 (29.5) | 81 (37.9) | 154 (33.0) | 0.262 |
| Aching | 19 (9.2) | 23 (10.7) | 46 (9.9) | 0.178 |
| Other* | 69 (33.3) | 72 (33.6) | 165 (35.4) | - |
| Pain Level on Initial<br>Consult |  |  |  |  |
| Mild Pain | 1 ( 0.5) | 14 ( 6.5) | 15 ( 3.2) | 0.334 |
| Moderate Pain | 10 ( 4.8) | 10 ( 4.7) | 20 ( 4.3) | 0.007 |
| Severe Pain | 100 (48.3) | 30 (14.0) | 133 (28.5) | 0.797 |
| Unilateral,<br>Sidelocked | 126 (60.9) | 106 (49.5) | 249 (53.4) | 0.231 |
| Nausea | 142 (68.6) | 72 (33.6) | 235 (50.4) | 0.776 |
| Vertigo | 7 (3.4) | 8 (3.7) | 19 (4.1) | 0.357 |
| Dizziness | 108 (52.2) | 62 (29.0) | 188 (40.3) | 0.561 |
| Vomiting | 52 (25.1) | 15 (7.0) | 73 (15.7) | 0.603 |
| Light Sensitivity | 176 (85.0) | 58 (27.1) | 260 (55.8) | 1.438 |
| Noise Sensitivity | 167 (80.7) | 53 (24.8) | 244 (52.4) | 1.355 |
| Visual Aura | 41 (19.8) | 17 (7.9) | 68 (14.6) | 0.481 |
| Tinnitus Onset | 0 (0.0) | 1 (0.5) | 1 (0.2) | 0.37 |
| Neck Pain | 96 (46.4) | 73 (34.1) | 185 (39.7) | 0.403 |
| Blurry Vision | 19 (9.2) | 8 (3.7) | 32 (6.9) | 0.411 |
| Osmophobia | 40 (19.3) | 8 (3.7) | 50 (10.7) | 0.603 |

Table 5 lists exacerbating or aggravating factors of the continuous baseline headache: stress, bright lights, flashing lights, or glare, loud noise, lack of sleep, strong smells, physical activity, change of weather, missed meals, exercise, certain foods, menses, alcohol, concentration, bending over, computer use, heat, reading, head jarring, and crying. The following were the five most reported exacerbating factors for people with MP: stress (8.7%), bright light, flashing light, or glare (23.2%), loud noise (24.2%), physical activity (23.7%), and exercise (8.7%). The four most reported aggravating causes for TP were, stress (10.3%), bright light, flashing light, or glare (5.6%), loud noise (5.6%), and physical activity (10.3%).

**Table 5.** Exacerbating Factors.

| Exacerbating Factor | Migraine Phenotype<br>Patients Seen<br>n=207 (%) | Tension Phenotype<br>Patients Seen<br>n=214 (%) | Total Number of<br>Patients Seen<br>n= 466 (%) | SMD |
| --- | --- | --- | --- | --- |
| Stress (%) | 18 (8.7) | 22 (10.3) | 43 (9.2) | 0.15 |
| Bright light, flashing light, or glare (%) | 48 (23.2) | 12 (5.6) | 71 (15.2) | 0.524 |
| Loud Noise (%) | 50 (24.2) | 12 (5.6) | 71 (15.2) | 0.545 |
| Lack of Sleep (%) | 15 (7.2) | 7 (3.3) | 25 (5.4) | 0.217 |
| Strong Smells (%) | 13 (6.3) | 1 (0.5) | 16 (3.4) | 0.348 |
| Physical Activity (%) | 49 (23.7) | 22 (10.3) | 73 (15.7) | 0.374 |
| Change of Weather (%) | 10 (4.8) | 7 (3.3) | 20 (4.3) | 0.152 |
| Missed Meals (%) | 2 (1.0) | 0 (0.0) | 2 (0.4) | 0.192 |
| Exercise (%) | 18 (8.7) | 7 (3.3) | 26 (5.6) | 0.259 |
| Certain Foods (%) | 3 (1.4) | 2 (0.9) | 5 (1.1) | 0.141 |
| Menses (%) | 8 (3.9) | 5 (2.3) | 13 (2.8) | 0.157 |
| Alcohol (%) | 3 (1.4) | 2 (0.9) | 5 (1.1) | 0.141 |
| Concentration (%) | 6 (2.9) | 6 (2.8) | 12 (2.6) | 0.134 |
| Bending Over (%) | 9 (4.3) | 5 (2.3) | 16 (3.4) | 0.171 |
| Computer Use (%) | 4 (1.9) | 4 (1.9) | 11 (2.4) | 0.134 |
| Heat (%) | 2 (1.0) | 1 (0.5) | 3 (0.6) | 0.146 |
| Reading (%) | 1 (0.5) | 1 (0.5) | 2 (0.4) | 0.134 |
| Head Jarring (%) | 1 (0.5) | 0 (0.0) | 1 (0.2) | 0.166 |
| Crying (%) | 2 (1.0) | 1 (0.5) | 4 (0.9) | 0.146 |

Table 6 provides the data on prognostic types for the persisting/refractory, (termed refractory by ICHD-3), relapsing–remitting, and remitting/self-limited with or without treatment for all subjects, MP, and TP. Table 7 provides the data on the number of patients with chronic headache lasting for 5 years or longer.

**Table 6.** Prognostic Type of Headache.

| Prognostic Type of Headache (%) | Migraine Phenotype<br>Patients Seen<br>n=207 (%) | Tension Phenotype<br>Patients Seen<br>n=214 (%) | Total Number of<br>Patients Seen<br>n= 466 (%) | SMD |
| --- | --- | --- | --- | --- |
| Persisting | 184 (88.9) | 201 (93.9) | 428 (91.8) | 0.208 |
| Relapsing remitting | 5 (2.4) | 5 (2.3) | 12 (2.6) |  |
| Remitting | 8 (3.9) | 4 (1.9) | 12 (2.6) |  |
| NA | 10 (4.8) | 4 (1.9) | 14 (3.0) |  |

**Table 7.** Years with Chronic Headache.

| Years with Chronic Headache | Migraine Phenotype Patients Seen<br>n=207 (%) | Tension Phenotype Patients Seen<br>n=214 (%) |
| --- | --- | --- |
| 5–10 years | 16.60% | 13.90% |
| 10–15 years | 7.50% | 5.30% |
| 15–20 years | 0.50% | 2.60% |
| 20–25 years | 0.50% | 0% |
| More than 25 years | 1% | 1.60% |

### Conclusions

NDPH is an enigmatic, treatment resistant condition with poorly understood etiology resulting in some patients suffering from years of continuous, unremitting pain. Contributing to this knowledge gap is the relative rarity of NDPH^16,22,23^, resulting in a potentially myopic view of a syndrome that is a diagnosis of exclusion. Therefore, a large-scale retrospective chart review of 466 cases was conducted deliberately aligning with the study design of one of the prior largest studies by Evans^21^ in order to compare and generalize findings of NDPH features.

The mean age of onset for this study was 37.1, which is consistent with other published works (28 to 40 years)^18,19,24^ and Evans (40.3 years) (Table 8). The observed data showed no obvious pattern in onset month similar to Evans. The racial and ethnic composition of our sample differed significantly from that of Evans, with our cohort predominantly identifying as White, whereas the Evans study included a substantially larger proportion of Hispanic and Black patients (Table 8). This study’s gender breakdown did not significantly differ from Evans with a predominance of females in, approximately, a 2:1 ratio (Table 8). The duration of NDPH symptoms at the last visit was similar to Evans, though this study found a statistically significant lower prior history of reported migraine.

**Table 8.** Ours vs Evans et al., 2021 Demographics.

| Demographic | Our Data, All Patients n=466 (%) | Evans et al., 2021<sup>21</sup> Data, All Patients (%) n=328 | P value |
| --- | --- | --- | --- |
| Age at onset, mean (SD) | 37.1 (18.6) | 40.3 (17.2) | <0.05 |
| Onset month, n (%) |  |  | NS |
| January | 40 (8.6) | 25 (7.6) |  |
| February | 24 ( 5.2) | 25 (7.6) |  |
| March | 20 ( 4.3) | 17 (5.2) |  |
| April | 33 ( 7.1) | 22 (6.7) |  |
| May | 17 ( 3.6) | 19 (5.8) |  |
| June | 18 ( 3.9) | 28 (8.5) |  |
| July | 23 ( 4.9) | 24 (7.3) |  |
| August | 16 ( 3.4) | 16 (4.9) |  |
| September | 34 ( 7.3) | 22 (6.7) |  |
| October | 35 ( 7.5) | 22 (6.7) |  |
| November | 24 ( 5.2) | 13 (4.0) |  |
| December | 23 ( 4.9) | 18 (5.5) |  |
| Unknown | 159 (34.1) | 77 (23.5) |  |
| Gender, n(%) |  |  | NS |
| Female | 311 (66.7) | 215 (65.5) |  |
| Male | 143 (30.7) | 113 (34.5) |  |
| Ethnicity, n (%) |  |  | <0.001 |
| Asian | 5 (1.1) | 10 (3.1) |  |
| Black | 35 (7.5) | 41 (12.5) |  |
| Hispanic | 3 (0.6) | 48 (14.6) |  |
| White | 400 (85.8) | 229 (69.8) |  |
| Other | 3 (0.6) | - |  |
| Unknown | 20 (4.29) | - |  |
| Duration of NDPH at last visit (years), median [25th, 75th] | 1.75 [0.58, 5] | 1.9 [0.7, 4.8] |  |
| History of migraine, n (%) | 48 (10.3) | 96 (29.3) | <0.001 |
| Initial Lateralization of Pain, n (%) |  |  |  |
| Unilateral (*Left or Right) | 56 (12.0) | - |  |
| Bilateral | 276 (59.2) | - |  |
| Left | 34 (7.3) | - |  |
| Right | 22 (4.7) | - |  |
| Unknown | 134 (28.8) | - |  |
| Final Lateralization of Pain (%) |  |  |  |
| Unilateral (*Left or Right) | 46 ( 9.9) | - |  |
| Bilateral | 318 (68.2) | - |  |
| Left | 27 ( 5.8) | - |  |
| Right | 19 ( 4.1) | - |  |
| Unknown | 102 (21.9) | - |  |

While stressful life events were reported as the primary precipitant in the Evans study, infection emerged as the most frequent trigger within our cohort, with both studies showing similar proportions of infection-related cases (Table 9). When comparing clinical features between both studies, there was a similar prevalence of thunderclap onset, nausea, vertigo, and vomiting. Our population had decreased prevalence of throbbing, pressure, aching, light sensitivity, and noise sensitivity, while having an increased prevalence of unilateral side-locked pain and visual aura. As for distribution of pain, our population mostly reported severe pain when such pain data was present, while Evans reported pain was mostly moderate (Table 10). For exacerbating factors, our population had increased reports of light sensitivity, sound sensitivity, physical activity and exercise, while also having lower reported rates of exacerbation by foods and missed meals (Table 11). Prognosis between both studies finds mostly a persisting subtype of NDPH with small populations of remitting, and relapsing remitting (Table 12).

**Table 9.**
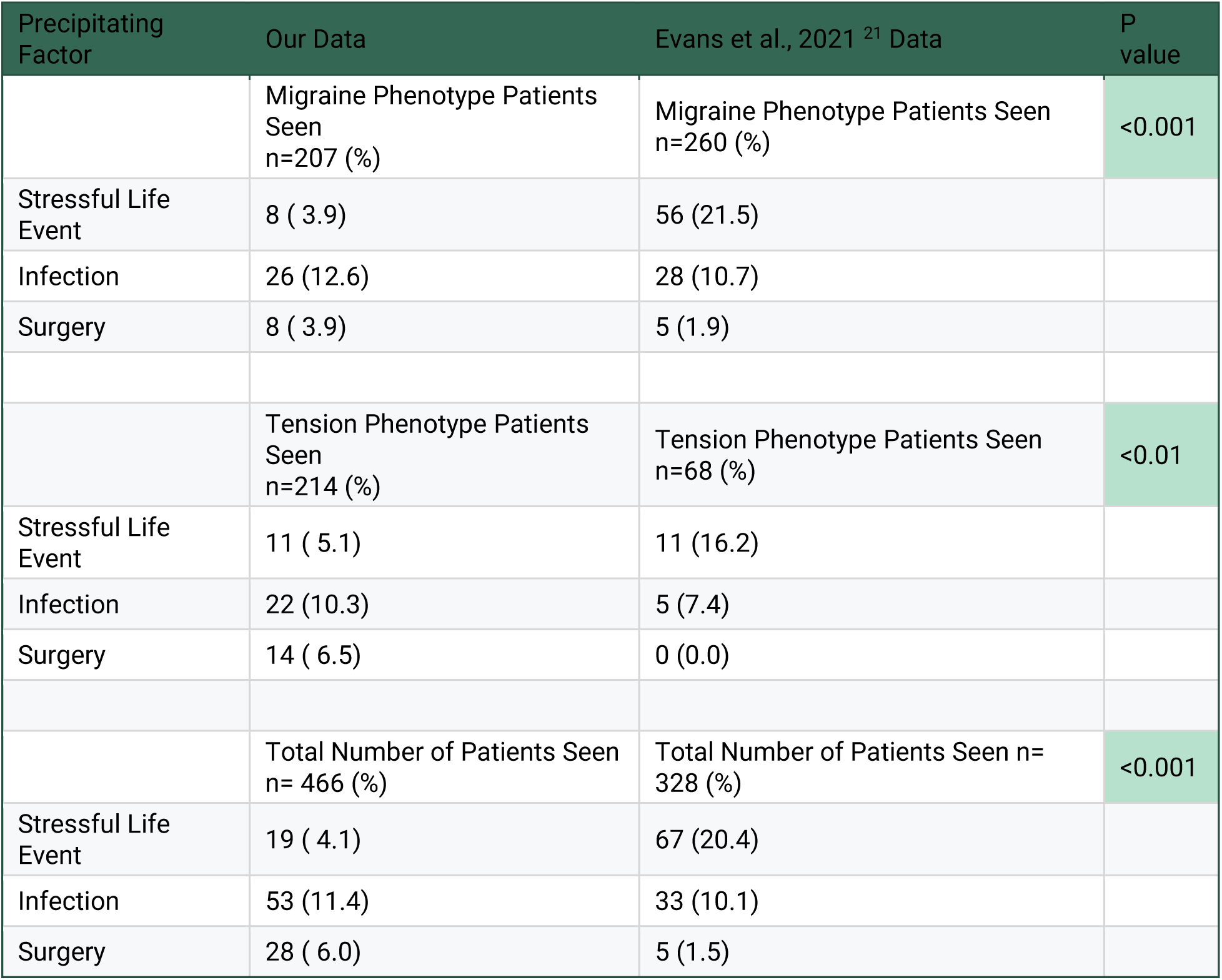
Ours vs Evans et al., 2021 Comparison of Precipitating Factors.

| Precipitating Factor | Our Data | Evans et al., 2021 <sup>21</sup> Data | P value |
| --- | --- | --- | --- |
|  | Migraine Phenotype Patients Seen<br>n=207 (%) | Migraine Phenotype Patients Seen<br>n=260 (%) | <0.001 |
| Stressful Life Event | 8 ( 3.9) | 56 (21.5) |  |
| Infection | 26 (12.6) | 28 (10.7) |  |
| Surgery | 8 ( 3.9) | 5 (1.9) |  |
|  | Tension Phenotype Patients Seen<br>n=214 (%) | Tension Phenotype Patients Seen<br>n=68 (%) | <0.01 |
| Stressful Life Event | 11 ( 5.1) | 11 (16.2) |  |
| Infection | 22 (10.3) | 5 (7.4) |  |
| Surgery | 14 ( 6.5) | 0 (0.0) |  |
|  | Total Number of Patients Seen<br>n= 466 (%) | Total Number of Patients Seen n= 328 (%) | <0.001 |
| Stressful Life Event | 19 ( 4.1) | 67 (20.4) |  |
| Infection | 53 (11.4) | 33 (10.1) |  |
| Surgery | 28 ( 6.0) | 5 (1.5) |  |

**Table 10.** Ours vs Evans et al., 2021 Comparison of Clinical Features.

| Clinical Features | Total Number of Patients<br>Seen<br>n= 466 (%) | Total Number of Patients<br>n= 328 (%), Evans et al.,<br>2021<sup>21</sup> | P<br>Value |
| --- | --- | --- | --- |
| Thunderclap Onset | 27 (5.8) | 12 (3.7) | NS |
| Quality of Pain |  |  |  |
| Throbbing | 144 (30.9) | 167 (50.9) | <0.001 |
| Pressure | 154 (33.0) | 180 (54.9) | <0.001 |
| Aching | 46 (9.9) | 51 (15.5) | <0.05 |
| Other* | 165 (35.4) | 96 (29.3) |  |
| Pain Level on Initial<br>Consult |  |  | <0.001 |
| Mild Pain | 15 ( 3.2) | 28 (8.5) |  |
| Moderate Pain | 20 ( 4.3) | 190 (57.9) |  |
| Severe Pain | 133 (28.5) | 102 (31.1) |  |
| Unilateral, Sidelocked | 249 (53.4) | 28 (8.5) | <0.001 |
| Nausea | 235 (50.4) | 157 (47.9) | NS |
| Vertigo | 19 (4.1) | 20 (6.1) | NS |
| Dizziness | 188 (40.3) | - |  |
| Vomiting | 73 (15.7) | 48 (14.6) | NS |
| Light Sensitivity | 260 (55.8) | 233 (71.0) | <0.001 |
| Noise Sensitivity | 244 (52.4) | 236 (72.1) | <0.001 |
| Visual Aura | 68 (14.6) | 21 (6.4) | <0.001 |
| Tinnitus Onset | 1 (0.2) | - |  |
| Neck Pain | 185 (39.7) | - |  |
| Blurry Vision | 32 (6.9) | - |  |
| Osmophobia | 50 (10.7) | - |  |

**Table 11.** Ours vs Evans et al., 2021 Comparison of Exacerbating Factors.

| Exacerbating Factor | Total Number of Patients<br>n= 466 (%) | Total Number of Patients n = 328 (%)<br>Evans et al.,2021<sup>21</sup> | P<br>value |
| --- | --- | --- | --- |
| Stress (%) | 43 (9.2) | 42 (12.8) | NS |
| Bright light, flashing light, or glare (%) | 71 (15.2) | 28 (8.5) | <0.01 |
| Loud Noise (%) | 71 (15.2) | 22 (6.7) | <0.001 |
| Lack of Sleep (%) | 25 (5.4) | 20 (6.1) | NS |
| Strong Smells (%) | 16 (3.4) | 13 (4.0) | NS |
| Physical Activity (%) | 73 (15.7) | 12 (3.7) | <0.001 |
| Change of Weather (%) | 20 (4.3) | 12 (3.7) | NS |
| Missed Meals (%) | 2 (0.4) | 9 (2.7) | <0.01 |
| Exercise (%) | 26 (5.6) | 8 (2.4) | <0.05 |
| Certain Foods (%) | 5 (1.1) | 10 (3.0) | <0.05 |
| Menses (%) | 13 (2.8) | 7 (2.1) | NS |
| Alcohol (%) | 5 (1.1) | 7 (2.1) | NS |
| Concentration (%) | 12 (2.6) | 6 (1.8) | NS |
| Bending Over (%) | 16 (3.4) | 6 (1.8) | NS |
| Computer Use (%) | 11 (2.4) | 6 (1.8) | NS |
| Heat (%) | 3 (0.6) | 7 (2.1) | NS |
| Reading (%) | 2 (0.4) | 2 (0.6) | NS |
| Head Jarring (%) | 1 (0.2) | 2 (0.6) | NS |
| Crying (%) | 4 (0.9) | 1 (0.3) | NS |

**Table 12.** Ours vs Evans et al., 2021 Comparison of Prognostic Type.

| Prognostic Type of Headache | Our Data | Evans et al., 2021<sup>21</sup> Data | P Value |
| --- | --- | --- | --- |
|  | Migraine Phenotype Patients Seen<br>n=207 (%) | Migraine Phenotype Patients Seen<br>n=260 (%) | NS |
| Persisting | 184 (88.9) | 244 (93.9) |  |
| Relapsing remitting | 5 (2.4) | 6 (2.3) |  |
| Remitting | 8 (3.9) | 10 (3.8) |  |
|  | Tension Phenotype Patients Seen<br>n=214 (%) | Tension Phenotype Patients Seen<br>n=68 (%) | NS |
| Persisting | 201 (93.9) | 61 (89.7) |  |
| Relapsing remitting | 5 (2.3) | 3 (4.4) |  |
| Remitting | 4 (1.9) | 4 (5.9) |  |
|  | Total Number of Patients Seen<br>n= 466 (%) | Total Number of Patients Seen<br>n= 328 (%) | NS |
| Persisting | 428 (91.8) | 305 (93.0) |  |
| Relapsing remitting | 12 (2.6) | 9 (2.7) |  |
| Remitting | 12 (2.6) | 14 (4.3) |  |

Next, populations were stratified into MP and TP and compared to Evans. Overall, similar results were found (Table 13 and Table 14). Both MP and TP subpopulations demonstrated a younger age of onset, no pattern in month of onset, a higher proportion of white individuals, and lower reported migraine history. Analysis of precipitating factors revealed infection as the most common trigger in both subgroups, whereas Evans found stressful life event as the most prevalent (Table 10). For clinical features, the MP population exhibited decreased throbbing, decreased pressure, increased pain severity, increased unilaterality, decreased noise sensitivity, and increased visual aura (Table 15). The TP population exhibited decreased pressure, decreased aching, increased severe pain, increased unilaterality, increased nausea, increased vomiting, increased light sensitivity, and increased noise sensitivity (Table 16). For MP exacerbating factors there were increased reports of light sensitivity, noise sensitivity, physical activity, and exercise (Table 17). For TP exacerbating factors there was only a report of increased physical activity (Table 18). No differences in prognostic types were identified (Table 13).

**Table 13.** Ours vs Evans et al., 2021 Comparison Demographics of Migraine Phenotype.

| Demographic | Our Data, Number of Migraine-Type NDPH Patients (%)<br>n=207 | Number of Migraine-Type NDPH Patients (%)<br>n=260, Evans et al., 2021<sup>21</sup> | P Value |
| --- | --- | --- | --- |
| Age at onset, mean (SD) | 34.7(17.1) | 38.9 (16.5) | <0.01 |
| Onset month, n (%) |  |  |  |
| January | 19 (9.2) | 21 (8.1) | NS |
| February | 14 (6.8) | 17 (6.5) |  |
| March | 12 (5.8) | 16 (6.2) |  |
| April | 11 (5.3) | 17 (6.5) |  |
| May | 6 (2.9) | 12 (4.6) |  |
| June | 11 (5.3) | 22 (8.5) |  |
| July | 7 (3.4) | 22 (8.5) |  |
| August | 8 (3.9) | 12 (4.6) |  |
| September | 20 (9.7) | 18 (6.9) |  |
| October | 14 (6.8) | 18 (6.9) |  |
| November | 12 (5.8) | 11 (4.2) |  |
| December | 9 (4.3) | 14 (5.4) |  |
| Unknown | 64 (30.9) | 60 (23.1) |  |
| Race/ethnicity, n (%) |  |  | <0.001 |
| Asian | 2 (1.0) | 7 (2.7) |  |
| Black | 22 (10.6) | 35 (13.5) |  |
| Hispanic | 2 (1.0) | 45 (17.3) |  |
| White | 173(83.6) | 173 (66.5) |  |
| Unknown | 3 (1.4) | - |  |
| Duration of NDPH at last visit (years), median [25th, 75th] | 1.63 [0.58, 4.73] | 1.8 [0.7, 4.8] |  |
| History of migraine, n (%) | 31 (15.0) | 85 (32.7) | <0.001 |
| Initial Lateralization of Pain, n (%) |  |  |  |
| Unilateral (Left or Right) | 30 (14.5) | - |  |
| Bilateral | 120 (58.0) | - |  |
| Left | 16 (7.7) | - |  |
| Right | 14 (6.8) | - |  |
| Unknown | 57 (27.5) | - |  |
| Final Lateralization of Pain (%) |  |  |  |
| Unilateral (Left or Right) | 28 (13.5) | - |  |
| Bilateral | 148 (71.5) | - |  |
| Left | 13 (6.3) | - |  |
| Right | 15 (7.2) | - |  |
| Unknown | 31 (15.0) | - |  |

**Table 14:** Ours vs Evans et al., 2021 Comparison Demographics of Tension Phenotype.

| Demographic | Our Data, Number of<br>Tension-Type NDPH Patients<br>(%)<br>n=214 | Number of Tension-Type NDPH Patients (%)<br>n=68, Evans et al.,<br>2021<sup>21</sup> | P<br>Value |
| --- | --- | --- | --- |
| Age at onset, mean (SD) | 39.2 (20.1) | 45.7 (18.7) | <0.05 |
| Onset month, n (%) |  |  | NS |
| January | 17 (7.9) | 4 (5.9) |  |
| February | 9 (4.2) | 8 (11.8) |  |
| March | 7 (3.3) | 1 (1.5) |  |
| April | 20 (9.3) | 5 (7.3) |  |
| May | 8 (3.7) | 7 (10.3) |  |
| June | 6 (2.8) | 6 (8.8) |  |
| July | 13 (6.1) | 2 (2.9) |  |
| August | 8 (3.7) | 4 (5.9) |  |
| September | 14 (6.5) | 4 (5.9) |  |
| October | 20 (9.3) | 4 (5.9) |  |
| November | 9 (4.2) | 2 (2.9) |  |
| December | 9 (4.2) | 4 (5.9) |  |
| Unknown | 74 (34.6) | 17 (25.0) |  |
| Race/ethnicity, n (%) |  |  | <0.05 |
| Asian | 3 (1.4) | 3 (4.4) |  |
| Black | 10 (4.7) | 6 (8.8) |  |
| Hispanic | 1 (0.5) | 3 (4.4) |  |
| White | 186 (86.9) | 56 (82.4) |  |
| Unknown | 0 (0.0) | - |  |
| Duration of NDPH at last visit<br>(years), median [25th, 75th] | 1.75 [0.58, 5.00] | 2.1 [0.7, 3.4] |  |
| History of migraine, n (%) | 13 ( 6.1) | 11 (16.2) | <0.05 |
| Initial Lateralization of Pain, n (%) |  |  |  |
| Unilateral (Left or Right) | 22 (10.2) | - |  |
| Bilateral | 138 (64.5) | - |  |
| Left | 14 (6.5) | - |  |
| Right | 8 (3.7) | - |  |
| Unknown | 54 (25.2) | - |  |
| Final Lateralization of Pain (%) |  |  |  |
| Unilateral (Left or Right) | 15 (7.0) | - |  |
| Bilateral | 137 (64.0) | - |  |
| Left | 11 (5.1) | - |  |
| Right | 4 (1.9) | - |  |
| Unknown | 62 (29.0) | - |  |

**Table 15.** Ours vs Evans 2021 Comparison of Clinical Features Among Migraine Phenotype.

| Clinical Features | Migraine Phenotype Patients<br>Seen<br>n=207 (%) | Migraine Phenotype Patients<br>Seen<br>n=260 (%), Evans et al., 2021<sup>21</sup> | P<br>Value |
| --- | --- | --- | --- |
| Thunderclap Onset | 14 (6.8) | 11 (4.2) | NS |
| Quality of Pain |  |  |  |
| Throbbing | 89 (43.0) | 144 (55.4) | <0.05 |
| Pressure | 61 (29.5) | 143 (55.0) | <0.001 |
| Aching | 19 (9.2) | 40 (15.4) | NS |
| Other* | 69 (33.3) | 76 (29.2) |  |
| Pain Level on Initial<br>Consult |  |  | <0.001 |
| Mild Pain | 1 ( 0.5) | 16 (6.2) |  |
| Moderate Pain | 10 ( 4.8) | 149 (57.3) |  |
| Severe Pain | 100 (48.3) | 91 (35.0) |  |
| Unilateral, Sidelocked | 126 (60.9) | 23 (8.8) | <0.001 |
| Nausea | 142 (68.6) | 157 (60.4) | NS |
| Vertigo | 7 (3.4) | 19 (7.3) | NS |
| Dizziness | 108 (52.2) | - |  |
| Vomiting | 52 (25.1) | 48 (18.5) | NS |
| Light Sensitivity | 176 (85.0) | 226 (86.9) | NS |
| Noise Sensitivity | 167 (80.7) | 229 (88.1) | <0.05 |
| Visual Aura | 41 (19.8) | 21 (8.1) | <0.001 |
| Tinnitus Onset | 0 (0.0) | - |  |
| Neck Pain | 96 (46.4) | - |  |
| Blurry Vision | 19 (9.2) | - |  |
| Osmophobia | 40 (19.3) | - |  |

**Table 16.**
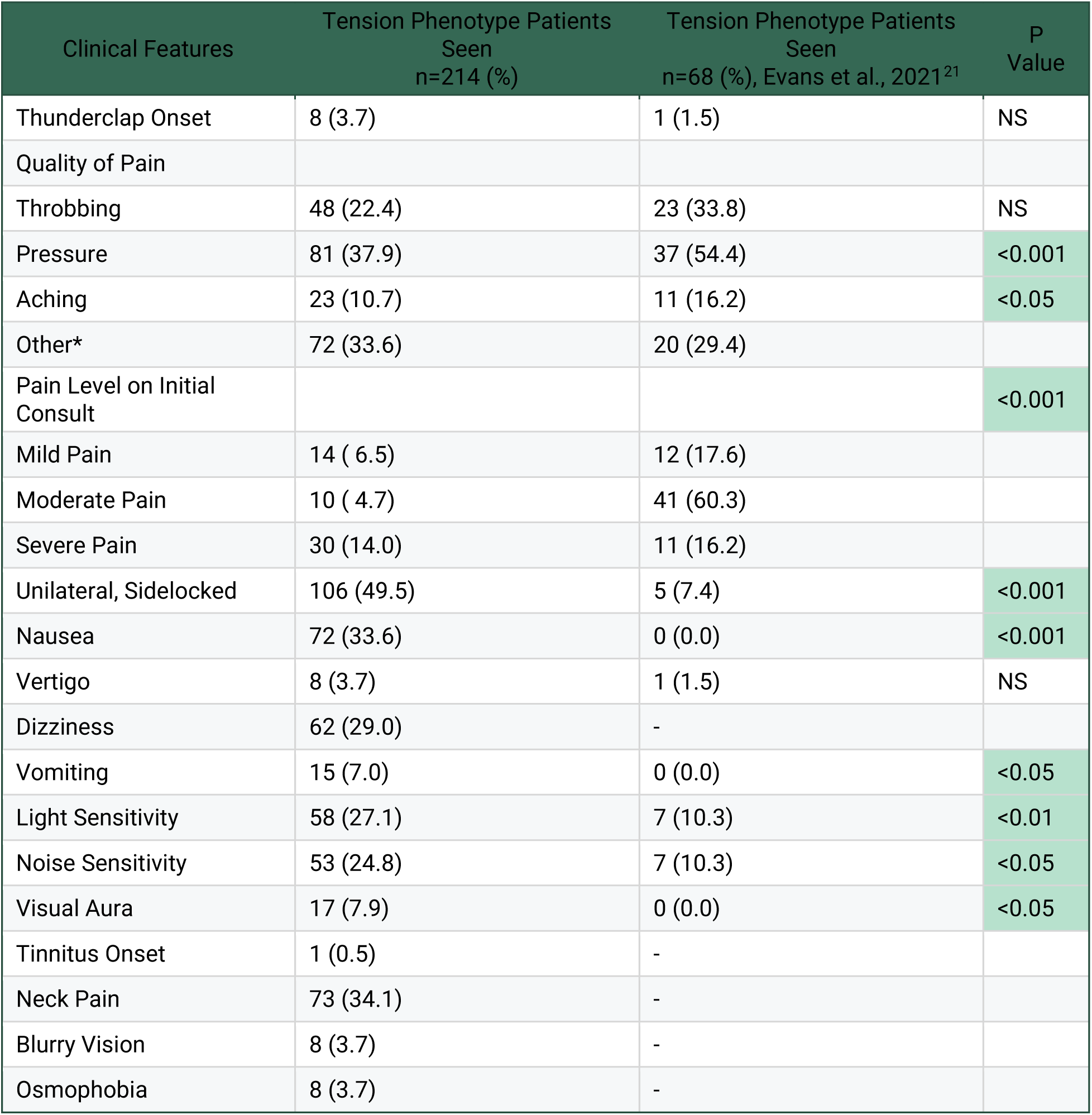
Ours vs Evans 2021 Comparison of Clinical Features Among Tension Phenotype.

**Table 17.** Ours vs Evans et al., 2021 Exacerbating Factors Among Migraine Phenotype.

| Exacerbating Factor | Migraine Phenotype Patients Seen<br>n=207 (%) | Migraine Phenotype Patients Seen<br>n=260 (%) , Evans et al., 2021<sup>21</sup> | P<br>value |
| --- | --- | --- | --- |
| Stress (%) | 18 (8.7) | 38 (14.6) | NS |
| Bright light, flashing light, or glare (%) | 48 (23.2) | 27 (10.4) | <0.001 |
| Loud Noise (%) | 50 (24.2) | 22 (8.5) | <0.001 |
| Lack of Sleep (%) | 15 (7.2) | 17 (6.5) | NS |
| Strong Smells (%) | 13 (6.3) | 13 (5.0) | NS |
| Physical Activity (%) | 49 (23.7) | 11 (4.2) | <0.001 |
| Change of Weather (%) | 10 (4.8) | 10 (3.8) | NS |
| Missed Meals (%) | 2 (1.0) | 9 (3.5) | NS |
| Exercise (%) | 18 (8.7) | 8 (3.1) | <0.01 |
| Certain Foods (%) | 3 (1.4) | 8 (3.1) | NS |
| Menses (%) | 8 (3.9) | 7 (2.7) | NS |
| Alcohol (%) | 3 (1.4) | 7 (2.7) | NS |
| Concentration (%) | 6 (2.9) | 6 (2.3) | NS |
| Bending Over (%) | 9 (4.3) | 6 (2.3) | NS |
| Computer Use (%) | 4 (1.9) | 5 (1.9) | NS |
| Heat (%) | 2 (1.0) | 5 (1.9) | NS |
| Reading (%) | 1 (0.5) | 2 (0.8) | NS |
| Head Jarring (%) | 1 (0.5) | 2 (0.8) | NS |
| Crying (%) | 2 (1.0) | 1 (0.4) | NS |

**Table 18.**
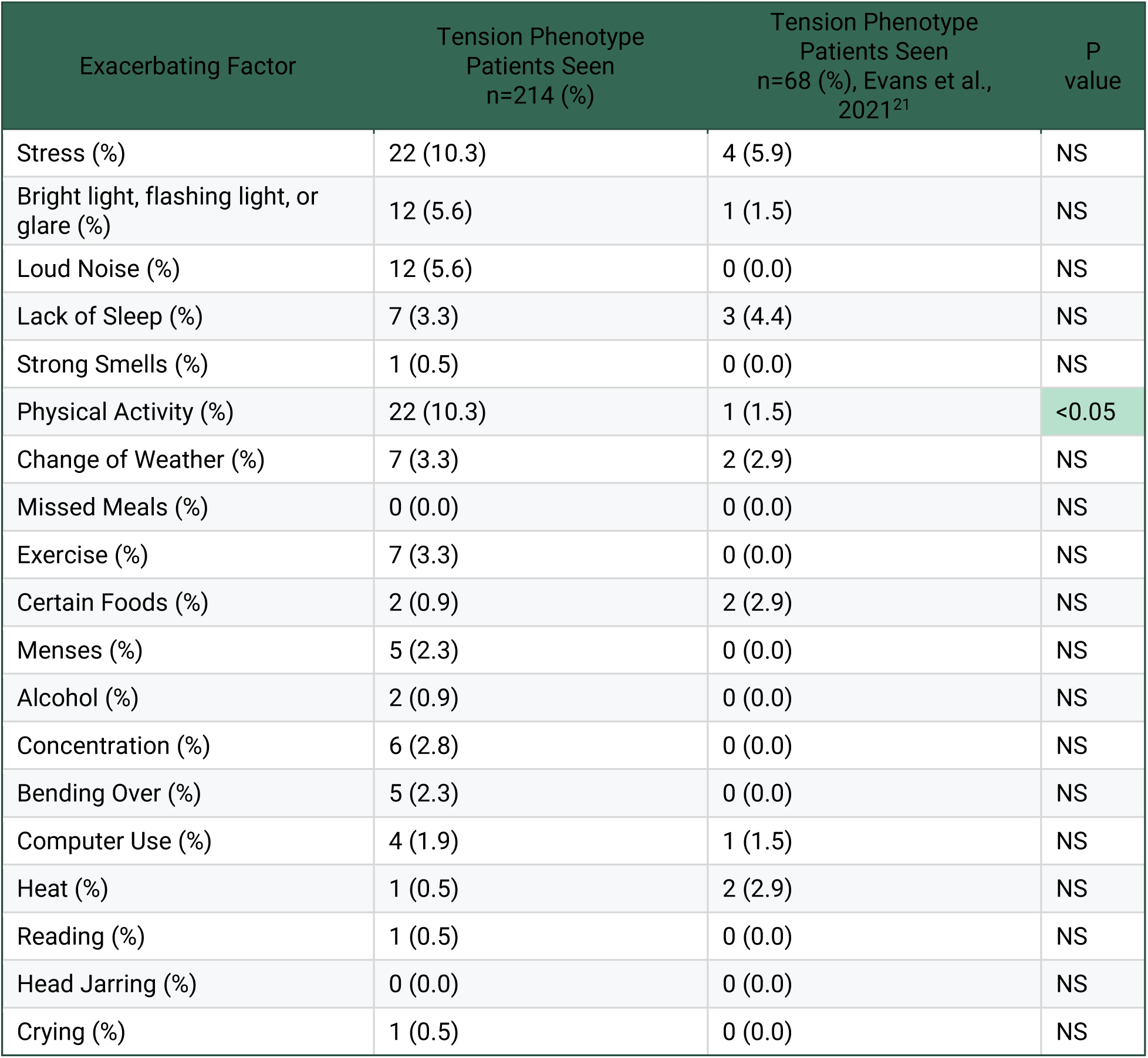
Ours vs Evans et al., 2021 Exacerbating Factors Among Tension Phenotype.

Overall, our study revealed numerous similarities with the findings reported by Evans. Gender breakdowns were similar, with an approximate 2:1 female-to-male ratio (Table 8). Both Evans and our work found no evidence of a seasonal pattern. Importantly, our study did not include juvenile patients, a population in which the month of headache onset has been linked to the beginning of the school year.^25^ It is possible that adult seasonality may arise in those who report an infectious triggering factor, given seasonal increases in viral infections.^26^ Although we looked specifically at NDPH patients who reported an infectious triggering factor, we did not observe any pattern in the month of onset (data not shown), though this may be due to a limited sample size.

An interesting observation within our population was significant rates of reported neck pain and exacerbation by physical activity, especially within our MP population (Table 4 and 5). This observation is interesting given the clinical overlap between NDPH and fibromyalgia. Both conditions share features such as joint hypermobility,^27,28^ viral association,^28,29^ TNF-alpha,^30–32^ COVID-19 association,^33,34^ headaches,^35,36^ and physical activity association.^37,38^ Importantly, according to the fibromyalgia diagnostic criteria, “a diagnosis of fibromyalgia is valid irrespective of other diagnoses. A diagnosis of fibromyalgia does not exclude the presence of other clinically important illnesses”.^39^ The association between migraine and fibromyalgia has also been well established.^40–42^ Considering this plausibility, we believe that further investigation would be appropriate to determine whether a fibromyalgia-associated subset of NPDH exists. Identifying such a subgroup may allow for more targeted treatment strategies, particularly with serotonin-norepinephrine reuptake inhibitors, which are first-line therapies for fibromyalgia and have shown promise in at least one reported NDPH case treated successfully with venlafaxine.^3^

Given the retrospective design of this work there is a significant limitation in interpretation. As noted in the introduction there is an interest in subtyping by triggering factor(s) in order to better identify treatments.^43^ However, between both Evans and our work the majority of patients did not have a reported triggering factor. It is possible that this is simply due to a failure to elicit that information or that reliance on triggering type subtyping may not be applicable in the majority of NDPH cases. Some of the focus on triggering type is due to reported treatment success in thunderclap onset patients^4^ and virally infected patients.^44^ Furthermore, while studies exist potentially pointing to disease etiologies they are limited due to study design, sample size, and/or failure to replicate, thereby limiting NDPH subtyping characteristics. Future work utilizing prospective follow up with particular focus on medication trial success and failures could greatly benefit and speed up identification of effective treatment.^21^

## Contributions

ZT concepted the study. ZT, RK designed the study. ZT, JM, BL, ND, VG, AV, SC, AC, FK, OB, AM, participated in data acquisition, analysis, and data interpretation. BL prepared final figures. ZT wrote the first draft of the manuscript. ZT, RK supervised all aspects of the work. All authors read and approved the final manuscript.

## Data Availability

Individualized patient data is not made available per IRB. Aggregated patient data is provided within.

## Declarations

### Ethics Approval

The University of Pittsburgh Institutional Review Board provided ethical approval and determined that the study was exempt and granted a waiver of HIPAA authorization. All work was performed in accordance with the 1964 Helsinki declaration and its later amendments.

### Consent for Publication

Not applicable.

## Acknowledgements

None.

## Rights and Permissions

Open Access This article is licensed under a Creative Commons Attribution-Non Commercial-No Derivatives 4.0 International License, which permits any non-commercial use, sharing, distribution and reproduction in any medium or format, as long as you give appropriate credit to the original author(s) and the source, provide a link to the Creative Commons license, and indicate if you modified the licensed material. You do not have permission under this license to share adapted material derived from this article or parts of it. The images or other third party material in this article are included in the article’s Creative Commons license, unless indicated otherwise in a credit line to the material. If material is not included in the article’s Creative Commons license and your intended use is not permitted by statutory regulation or exceeds the permitted use, you will need to obtain permission directly from the copyright holder. To view a copy of this license, visit http://creativecommons.org/licenses/by-nc-nd/4.0/.

## Abbreviation List

NDPH: New Daily Persistent Headache
ICD: International Classification of Diseases
UPMC: University of Pittsburgh, Medical Center System
ICHD-3: International Classification of Headache Disorders, 3rd edition
SD: Standard Deviations
SMD: Standardized Mean Differences
MP: Migraine Phenotype
TP: Tension-Type Phenotype

